# Inference of malaria transmission dynamics under varying assumptions about the spatial scale of transmission: a modelling study in three elimination settings

**DOI:** 10.1101/2020.09.27.20202630

**Authors:** Isobel Routledge, Samir Bhatt

## Abstract

**Background:** Individual-level geographic information about malaria cases, such as the GPS coordinates of residence or health facility, is often collected as part of surveillance in near-elimination settings, but could be more effectively utilised to infer transmission dynamics, in conjunction with additional information such as symptom onset time and genetic distance. However, in the absence of data about the flow of parasites between populations, the spatial scale of malaria transmission is often not clear. As a result, it is important to understand the impact of varying assumptions about the spatial scale of transmission on key metrics of malaria transmission, such as reproduction numbers.

**Methods:** We developed a method which allows the flexible integration of distance metrics (such as Euclidian distance, genetic distance or accessibility matrices) with temporal information into a single inference framework to infer malaria reproduction numbers. Twelve scenarios were defined, representing different assumptions about the likelihood of transmission occurring over different geographic distances and likelihood of missing infections (as well as high and low amounts of uncertainty in this estimate). These scenarios were applied to four individual level datasets from malaria eliminating contexts to estimate individual reproduction numbers and how they varied over space and time.

**Results:** Model comparison suggested that including spatial information improved models as measured by ΔAICc, compared to time only results. Across scenarios and across datasets, including spatial information tended to increase the seasonality of temporal patterns in reproduction numbers and reduced noise in the temporal distribution of reproduction numbers. The best performing parameterisations assumed long-range transmission (>200km) was possible.

**Conclusions:** Our approach is flexible and provides the potential to incorporate other sources of information which can be converted into distance or adjacency matrices such as travel times or molecular markers.

## Introduction

Individual-level disease surveillance data, collected routinely and as part of outbreak response, capture a wealth of information which could improve measurements of transmission and its spatiotemporal variation, in turn informing the design of epidemiological interventions^1^. Geo-located health facility or residence data are increasingly collected as part of surveillance for diseases such as malaria^2^, but could be more effectively utilised to infer transmission dynamics, in conjunction with additional information such as symptom onset time and genetic distance^3^. However, challenges exist in making use of these diverse data sources and leveraging the information they contain within a single inference framework. This is particularly true of endemic diseases such as malaria, where individual level data are increasingly collected in moderate-low transmission or elimination settings.

Malaria transmission is shaped by processes occurring on a wide range of spatial scales. In the absence of human mobility, transmission is limited to the range of the mosquito vector, however human movement, ranging from regular commutes to rare large scale migration events, can import parasites into new areas provided competent vectors are present^4–7^. As a result, the spatial location of individual cases can provide useful information in inferring transmission dynamics when combined with additional forms of information, such as temporal and molecular data. Furthermore, in elimination settings malaria transmission is thought to take on epidemic dynamics^8^, meaning the importance of space and highly dynamic factors such as human movement patterns becomes more relevant. However, in the absence of data about the flow of parasites between populations, the spatial scale of malaria transmission is often not clear. As a result, it is important to understand the impact of varying assumptions about the spatial scale of transmission on key metrics of malaria transmission, such as reproduction numbers. In many contexts, not all cases may be observed within a surveillance system. Missing cases further complicate inference, as without information about how likely cases are to be missing, ambiguity can exist as to whether long-range transmission occurred or whether cases were infected by a closer, unobserved source of infection.

To explore the impact of including distance measure and assumptions about their relationship to transmission likelihood, we developed a flexible framework to incorporate pairwise distances (for example Euclidian distances, travel times, or any quantifiable distance matrix) into our previously published inference framework^9^ to estimate individual reproduction numbers and explore the impact of varying assumptions about missing cases and the spatial kernel on results, as well as determining the feasibility of inferring the distance kernel and amount of missing cases from surveillance data. We defined twelve scenarios representing different assumptions about the likelihood of transmission occurring over different geographic distances and likelihood of missing infections (as well as high and low amounts of uncertainty in this estimate). These scenarios were applied to four individual level datasets from malaria eliminating contexts to estimate individual reproduction numbers and how they varied over space and time. We used two simple spatial kernels describing the relationship between Euclidian distance between residences and likelihood of transmission occurring, to explore various assumptions about the relationship between locations of cases and likelihood of transmission occurring between them, as well as the impact of unobserved cases. We find the best performing models by second order AIC (ΔAICc)^10^ have weakly informative priors on the likelihood of unobserved sources of infection and assume. However, we find there can be issues of parameter identifiability, which become increasingly relevant when there are not enough data available about key parameters in the model.

## Results

We developed a framework to integrate distance information into a previously published inference framework ^9,11^ which uses the time of symptom onset to infer reproduction numbers and their spatiotemporal variation. We then tested the impact of varying assumptions about the relationship between location of cases and the likelihood of transmission as well as the impact of unobserved infection as modelled by competing edges, ε, considering twelve scenarios (**Table 1**), and applying them to four line-list datasets from China (*P. vivax* and *P. falciparum*, analysed separately), El Salvador (*P. vivax*) and Eswatini (*P. falciparum*) using Exponential and Gaussian spatial kernels, described in the methods section of this paper.

**Table 1:**
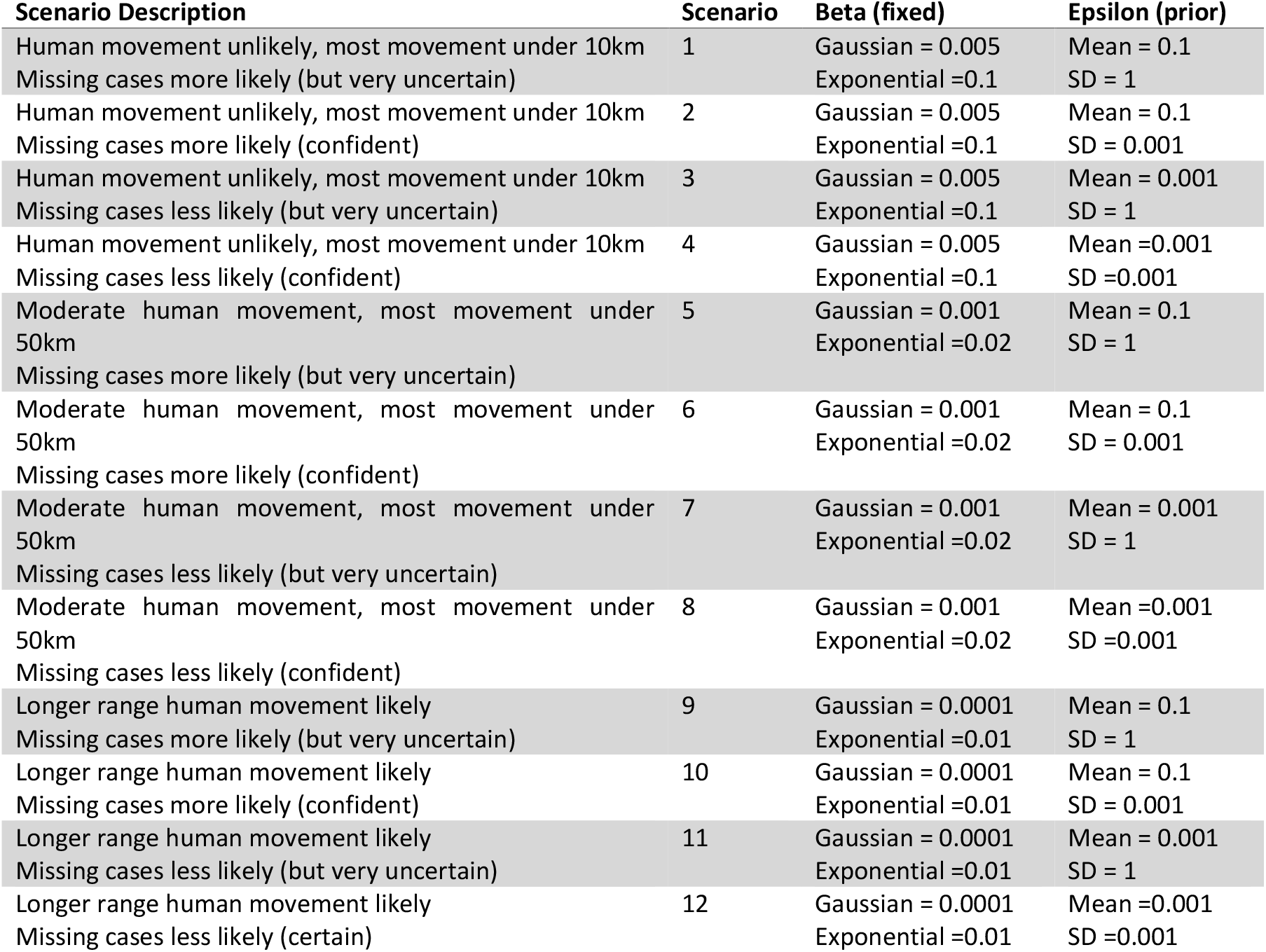
Table illustrating the different scenarios and corresponding parameter values tested in scenario analysis.

### Results of model comparison by ΔAICc across different scenarios

When ΔAICc scores were used to compare model results, all models which included distance had lower (and therefore better) ΔAICc scores than models which only included only time (**Table 2 and Supplementary Table 1**). In addition, exponential kernels consistently outperformed equivalent scenarios using Gaussian kernels **(Supplementary Table 1**). Two scenarios consistently performed best as measured by ΔAICc, namely Scenario 9 (El Salvador and Swaziland) and Scenario 11 (China, *P. vivax* and *P. falciparum*). Both scenarios assume longer range human movement likely and impose a smaller penalty on cases occurring larger distances. These scenarios also allow variation in epsilon edge values and use a very weakly informative prior on Epsilon edges, but with a different mean (0.1 for Scenario 9, 0.001 for Scenario 11). These results also return smaller mean *R*_*c*_ results than time-only versions of the model (**Figures 1–4**)

**Table 2:** Summary of ΔAICc results.

| Dataset | Best Model(s), by $\Delta AICc$ | Akaike Weight |
| --- | --- | --- |
| Swaziland (Eswatini) | Scenario 9, Exponential | 1 |
| El Salvador | Scenario 9, Exponential | 0.621540909785805 |
|  | Scenario 11, Exponential | 0.37845909 |
| China *P. vivax* | Scenario 11, Exponential | 1 |
| China *P. falciparum* | Scenario 11, Exponential | 1 |

**Figure 1:**
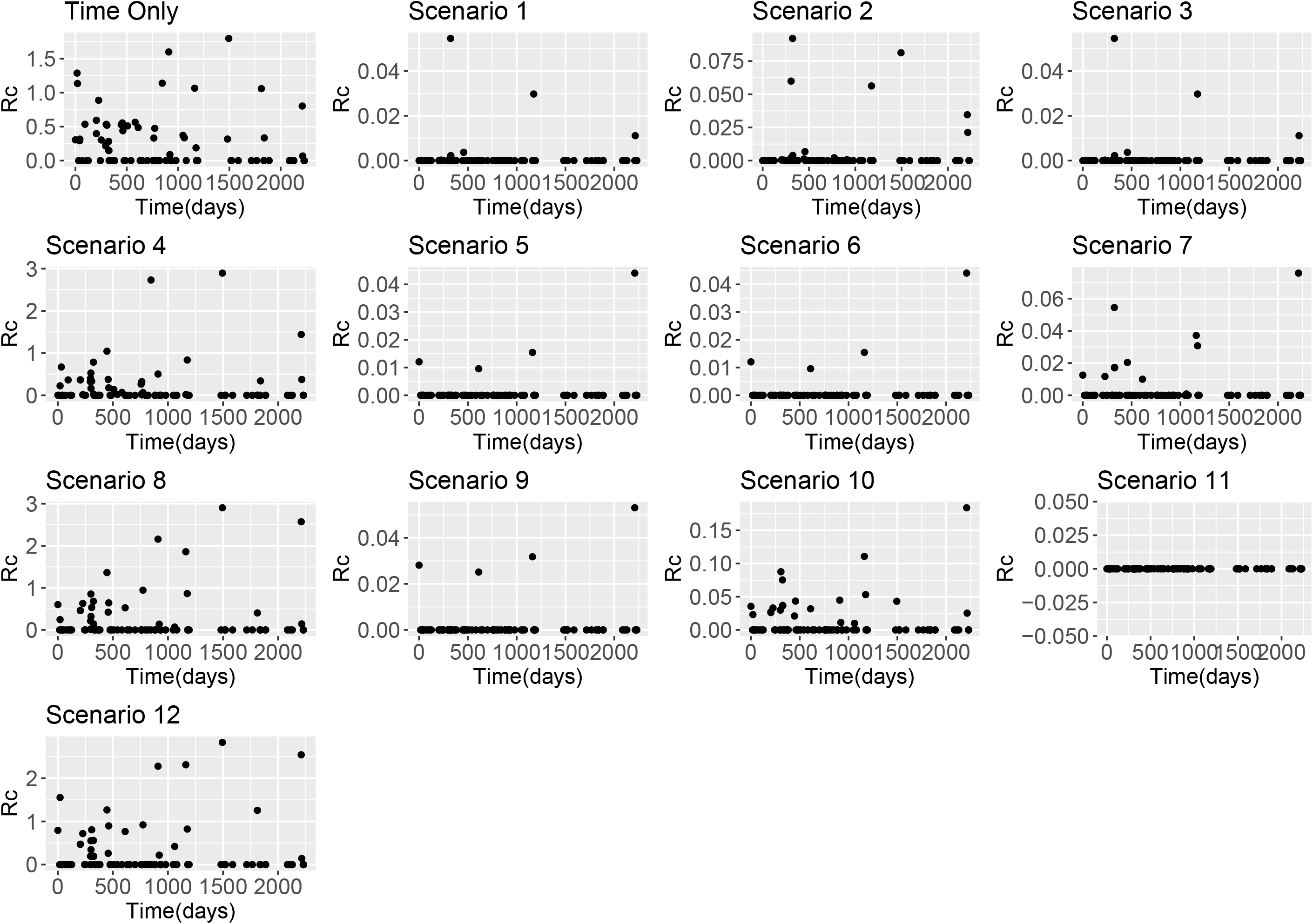
*R*_*c*_ estimates from El Salvador line list based on using the time-only scenario and Scenarios 1-12 with an exponential kernel

**Figure 2:**
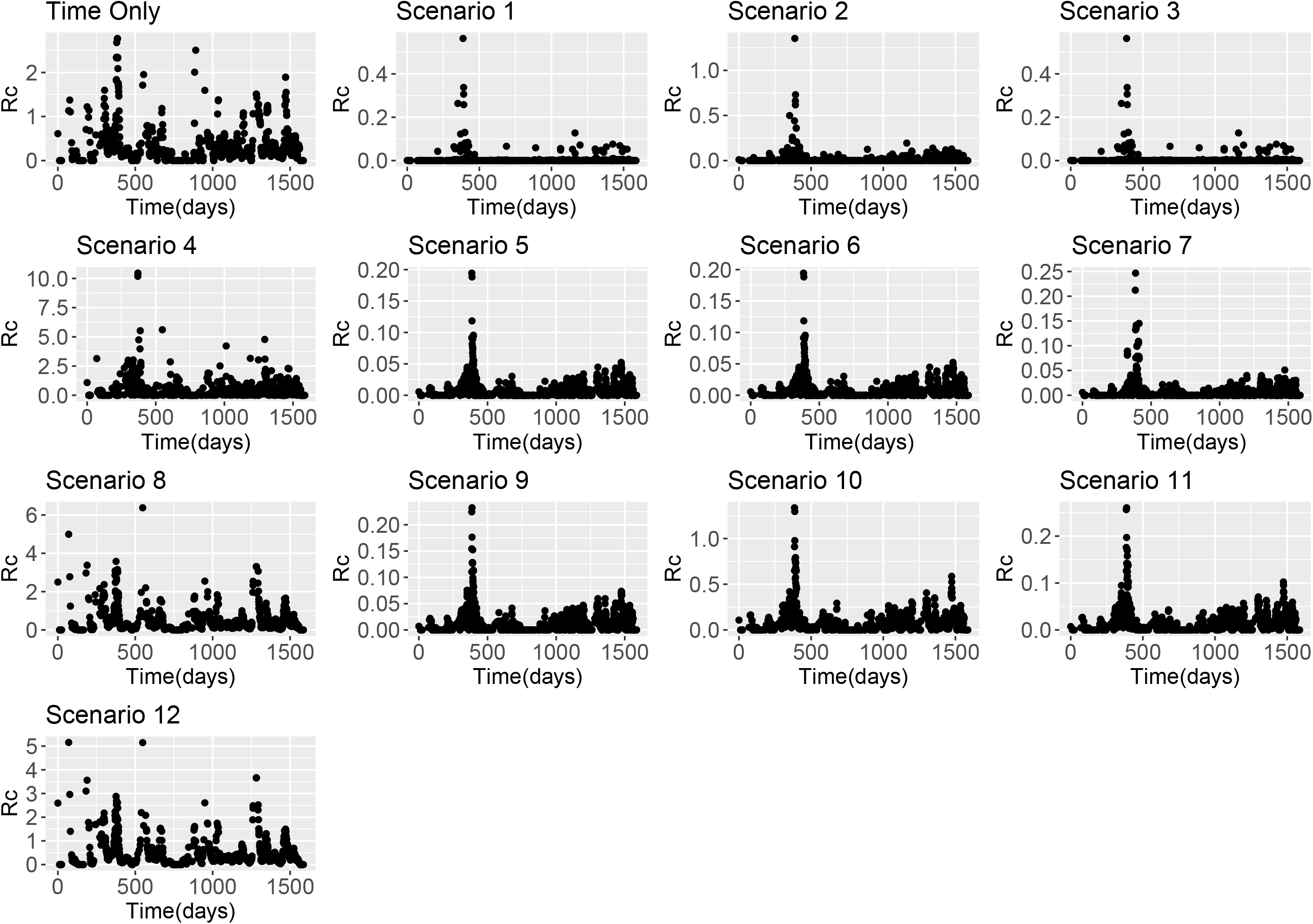
*R*_*c*_ estimates from Eswatini line list based on using the time-only scenario and Scenarios 1-12 with an exponential kernel

**Figure 3:**
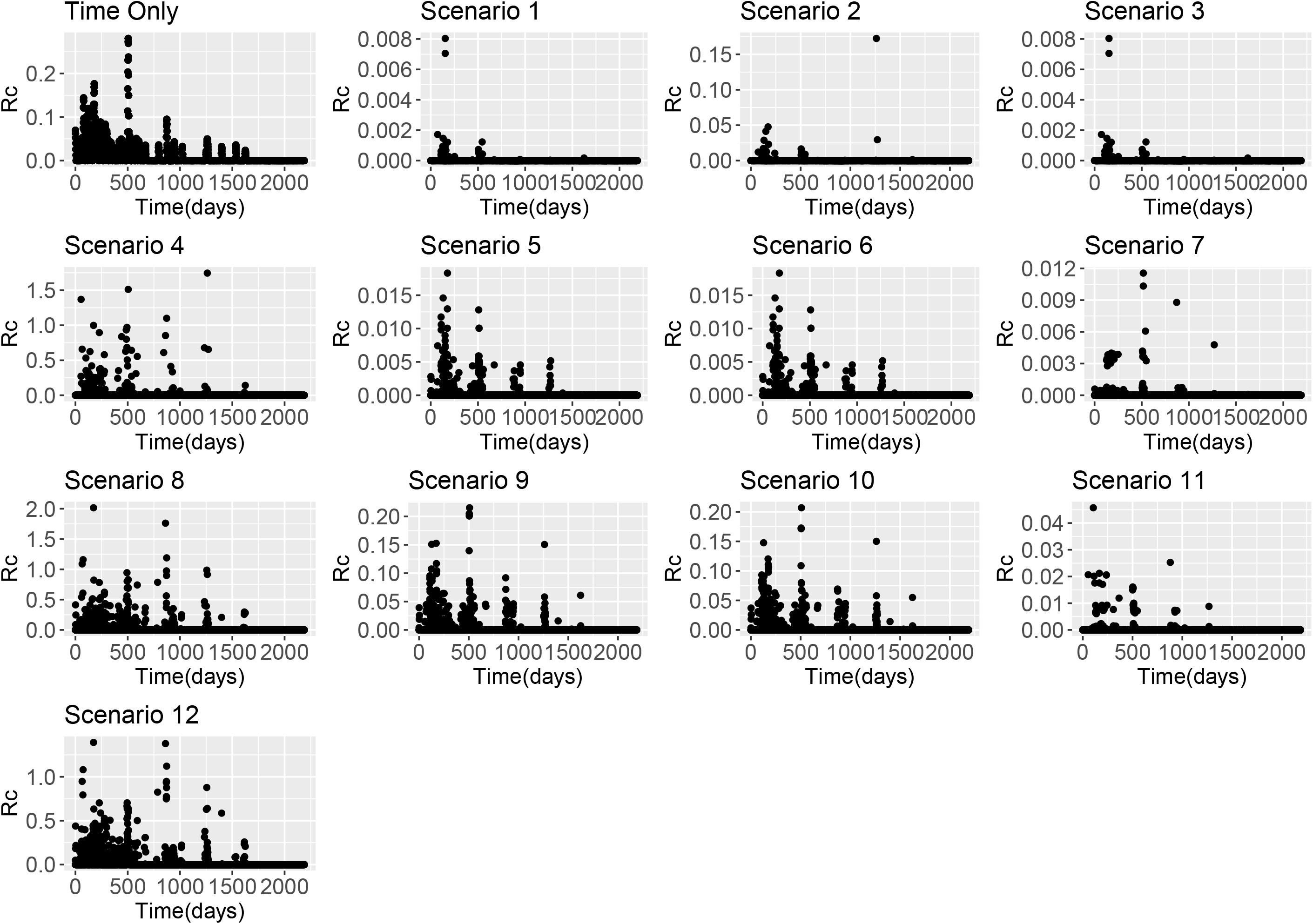
*R*_*c*_ estimates from China *P. falciparum* line list based on using the time-only scenario and Scenarios 1-12 with an exponential kernel

**Figure 4:**
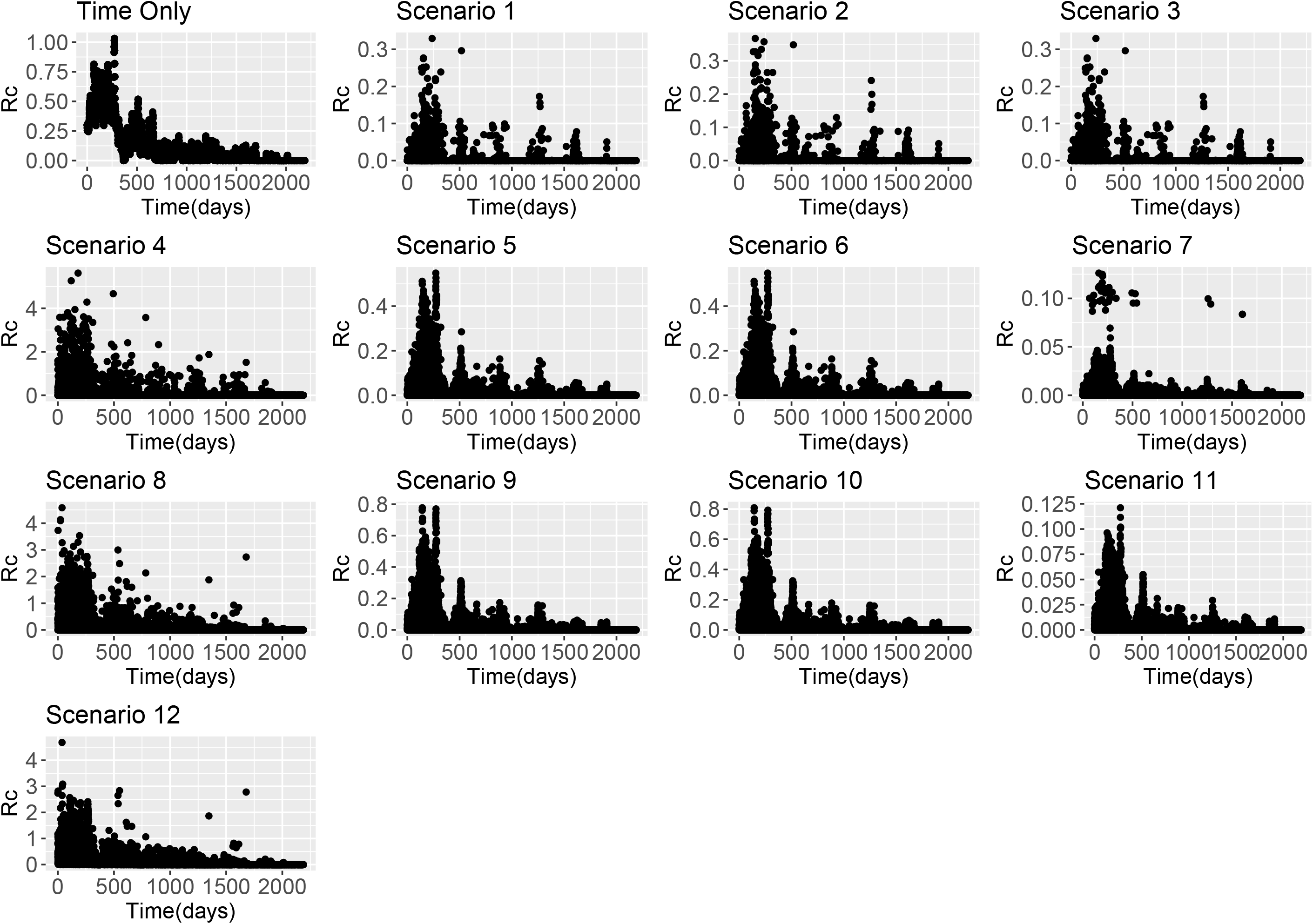
*R*_*c*_ estimates from China *P. vivax* line list based on using the time-only scenario and Scenarios 1-12 with an exponential kernel

### ***R***_***c***_ estimates under different scenarios

Across all datasets, large differences in estimates were found depending on both ε and β parameters. When β is higher, the assumption is that there is little movement of parasites within the country and therefore cases with residential addresses which are far away are unlikely to have infected each other. When this is the case and we assume there are unobserved sources of infection (either through a strongly informative prior on ε with mean 0.1, or an uninformative prior with a lower mean), then *R*_*c*_ values are very low. However if we assume there are little or no unobserved sources of infection, but continue to make restrictive assumptions about space, then most *R*_*c*_ very low but in the localities where there are cases we estimate much higher *R*_*c*_ values as there are no other possible infectors within a reasonable time and/or spatial area. This is illustrated in Figures 1 - 4.

When looking at the spatial patterns of *R*_*c*_ estimates under different scenarios several trends are seen across all datasets (**Figures 5-8**). Scenario 4 is particularly interesting to note because this scenario considers the most restrictive assumptions, both about space and unobserved sources of infection. Across datasets, Scenario 4 results in increased focality and higher *R*_*c*_s within these foci, but in comparison lower *R*_*c*_s in other areas. All of the best scenarios as measured by ΔAICc resulted in small *R*_*c*_ estimates, but where comparably larger *R*_*c*_ estimates were estimated, they were in localities identified as foci.

**Figure 5:**
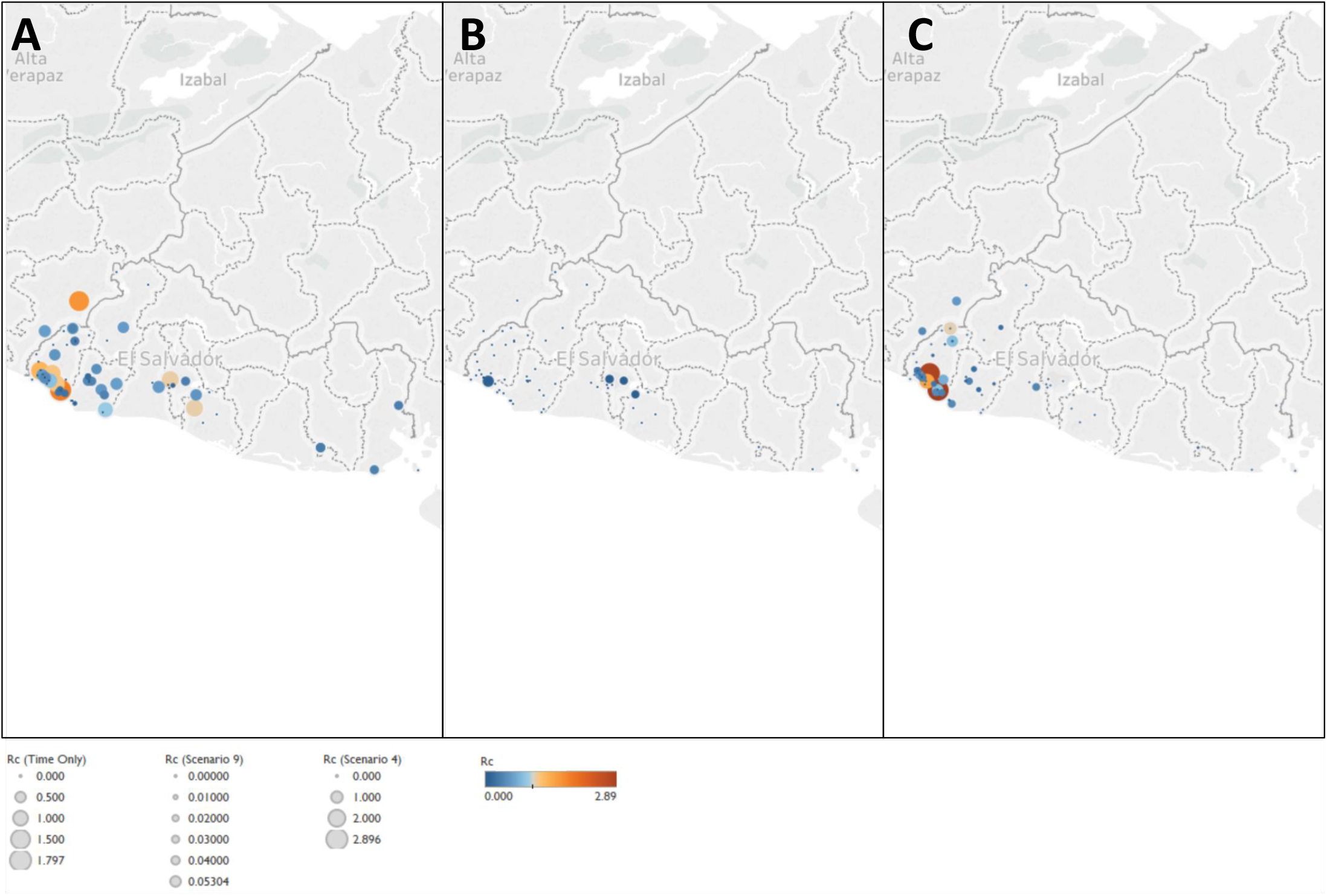
Map of Rc estimates for El Salvador. Map of A) Time-only B) Best scenario by AIC (Scenario 9) and C) Scenario 4, representing an assumption of little long-distance transmission and few unobserved cases. Note the increasing focality in C), with higher Rc values estimated on the Pacific Coastal area of the Ahuacapan and Sonsonote municipalities, where the NMCP have long identified as the remaining foci of risk.

**Figure 6:**
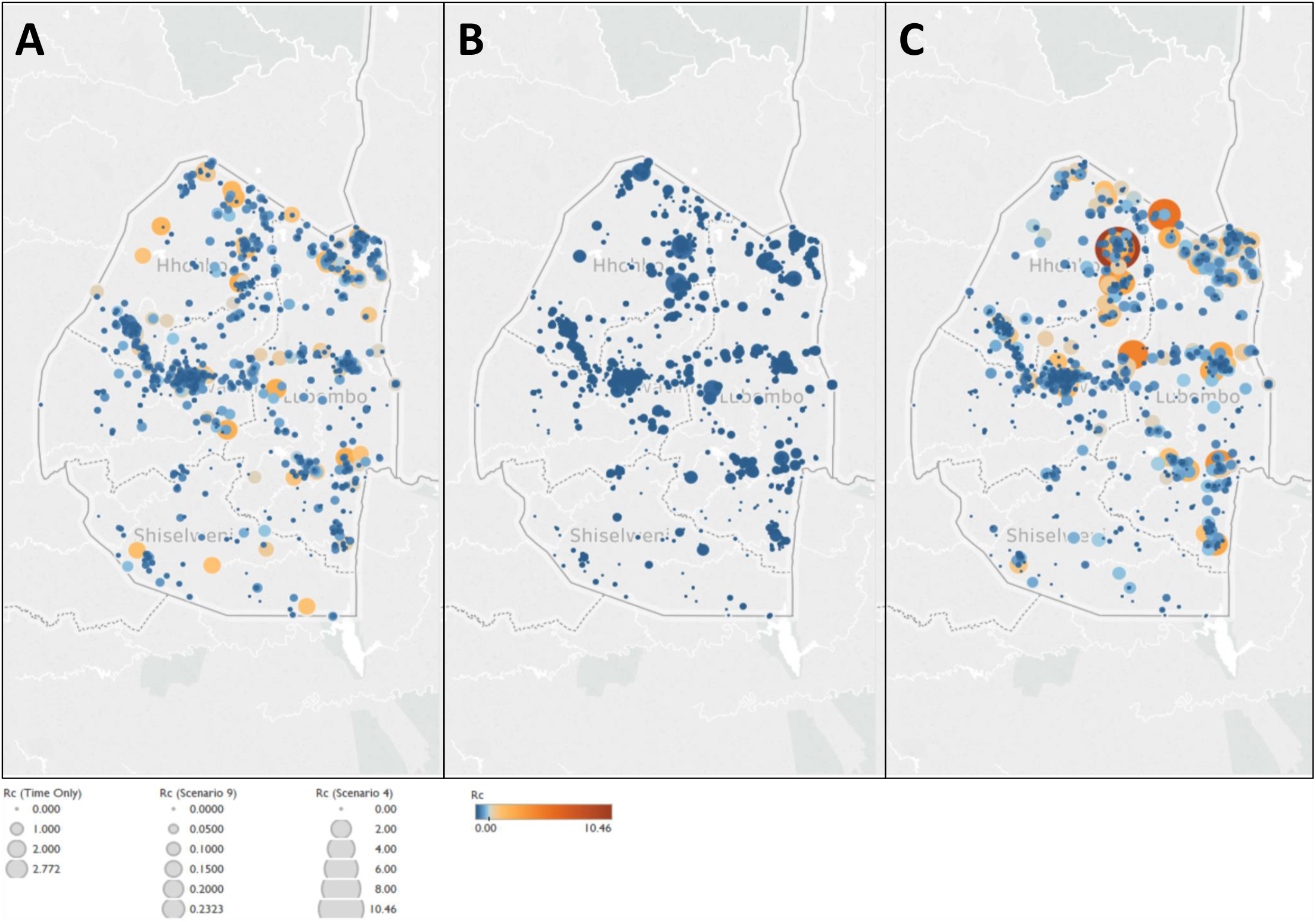
Map of Rc estimates for Swaziland. Map of A) Time-only B) Best scenario by AIC (Scenario 9) and C) Scenario 4, representing an assumption of little long-distance transmission and few unobserved cases. Note the increasing focality in C), with higher Rc values estimated around the northern corner of the country which borders Mozambique.

**Figure 7:**
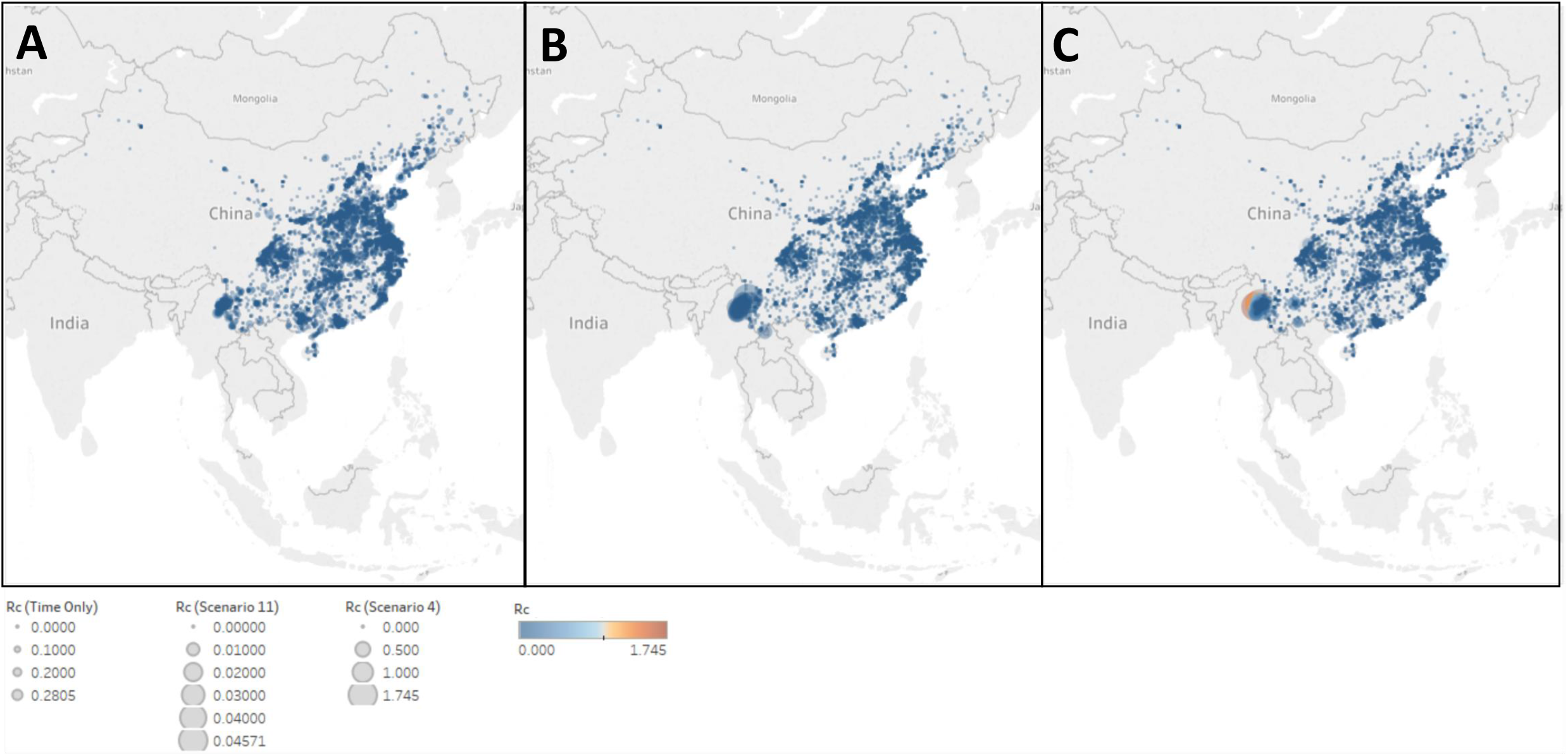
Map of Rc estimates for *P. falciparum* in China. Map of A) Time-only B) Best scenario by AIC (Scenario 11) and C) Scenario 4, representing an assumption of little long-distance transmission and few unobserved cases.

**Figure 8:**
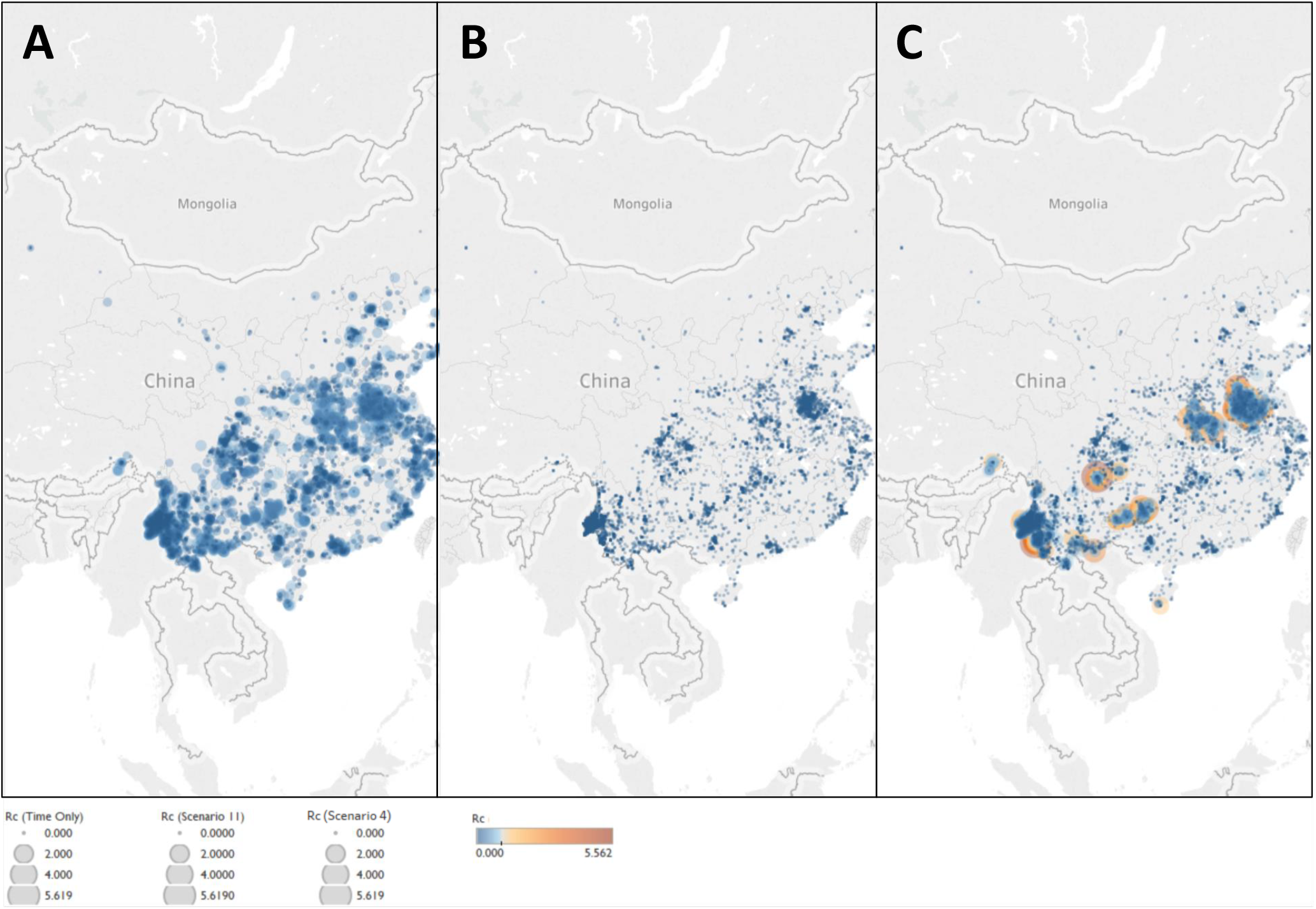
Map of Rc estimates for *P. vivax* in China. Map of A) Time-only B) Best scenario by AIC (Scenario 11) and C) Scenario 4, representing an assumption of little long-distance transmission and few unobserved cases.

For the line-list dataset from El Salvador, within the range of values explored in the sensitivity analysis (**Table 3**), regardless of how informative the prior was for either β, the distance shaping function, or for ε, the epsilon edge, β was always estimated as whatever the mean of the prior was set as between the prior mean values of 1e-4 and 1e-2 (**Figure 9**). However, when the mean value was set at 0.1, the estimated parameter converged at a slightly lower value of 0.075, with the exception of when the prior for ε was very low (all priors with mean ε of 1e-10 and also the more informative priors with mean 1e-5, when standard deviation was 1e-4). *R*_*c*_ is strongly shaped by the value of ε, with higher values of ε returning lower values of *R*_*c*_, however *R*_*c*_ also declined with increasing values of β.

**Table 3:** Different parameters considered in sensitivity analysis. Note all combinations of each parameter were considered.

| $\epsilon$ mean | $\epsilon$ SD | $\beta$ mean<br>(Gaussian) | $\beta$ mean<br>(Exponential) | $\beta$ SD |
| --- | --- | --- | --- | --- |
| **1e-10** | 0.0001 | 0.00001 | 0.0001 | 0.0001 |
| **1e-5** | 0.001 | 0.0001 | 0.001 | 0.001 |
| **1e-3** | 0.01 | 0.001 | 0.01 | 0.01 |
| **1e-2** | 0.05 | 0.01 | 0.1 | 0.05 |
| **1e-1** | 0.1 |  |  | 0.1 |
| **0.5** |  |  |  |  |

**Figure 9:**
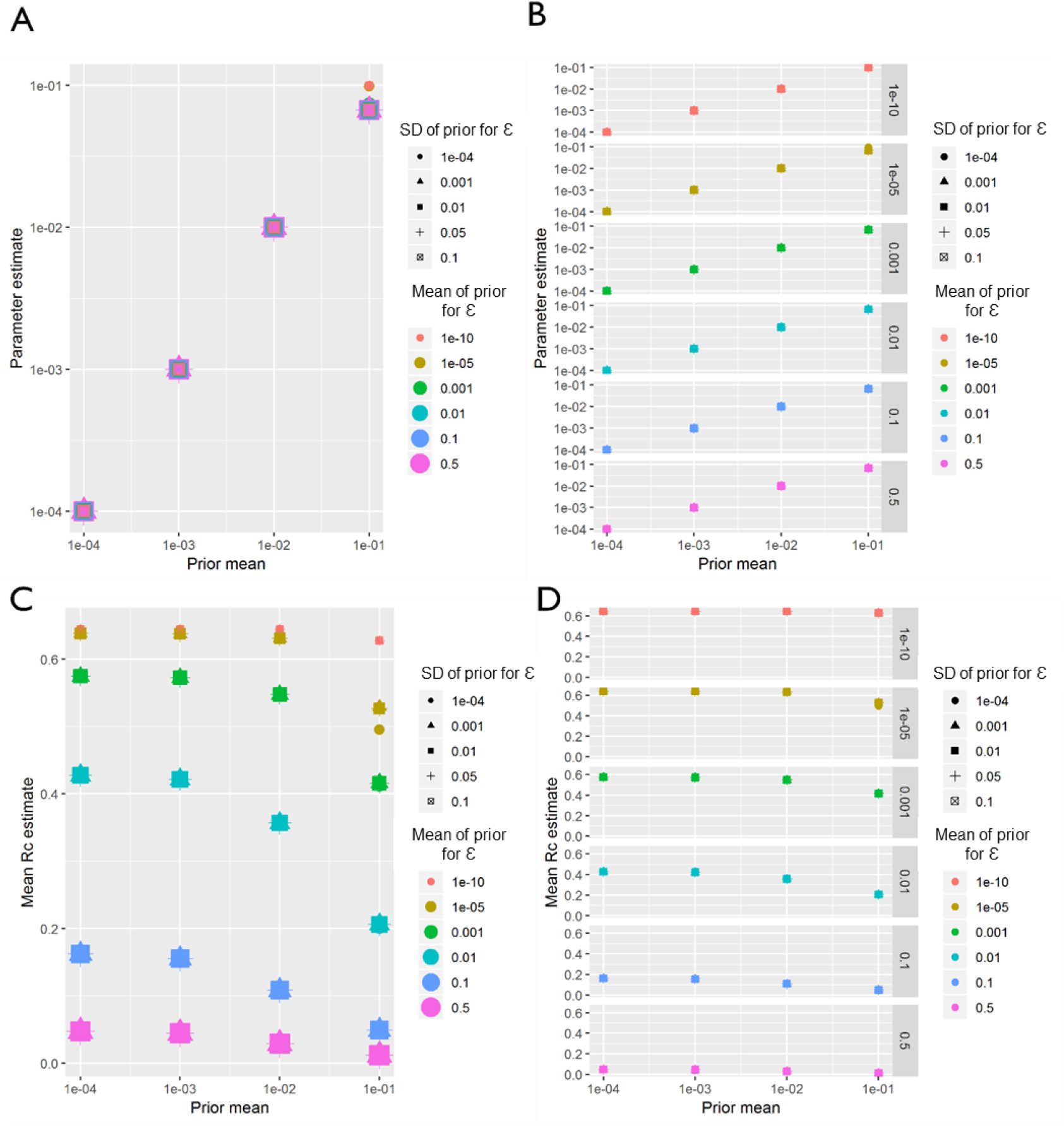
El Salvador sensitivity analysis. Sensitivity analysis showing the impact of varying the prior mean for the distance kernel shaping parameter, β. The different colours and shapes represent different means and standard deviations respectively of the normally-distributed prior of epsilon, ε, which represents shapes represent different hazards of infection by an external, unobserved source. For A-D, the x-axis represents the prior mean used for β. A) the y-axis shows the maximum a posteriori parameter estimate for the parameter β. B) shows the same results, stratified by the prior mean of ε for clarity. C) Shows the impact of priors for β and ε on the mean Rc estimate, and again D) shows the same result, stratified by the prior mean of ε.

Very similar patterns to El Salvador were observed in the sensitivity analysis of the Eswatini dataset. Again, regardless of how informative the prior was for either ε or β, β was always estimated as whatever the mean of the prior was set as between the prior mean values of 1e-4 and 1e-2 **(Figure 10**). However, when the mean value was set at 0.1, the estimated parameter converged at a slightly lower value of 0.075, with the exception of when the prior for ε was very low (all priors with mean ε of 1e-10 and also the more informative priors with mean 1e-5, when standard deviation was 1e-4). Unlike El Salvador, for Eswatini, at higher values of ε (0.5 and 0.1) there are stark declines in *R*_*c*_ with increasing β.

**Figure 10:**
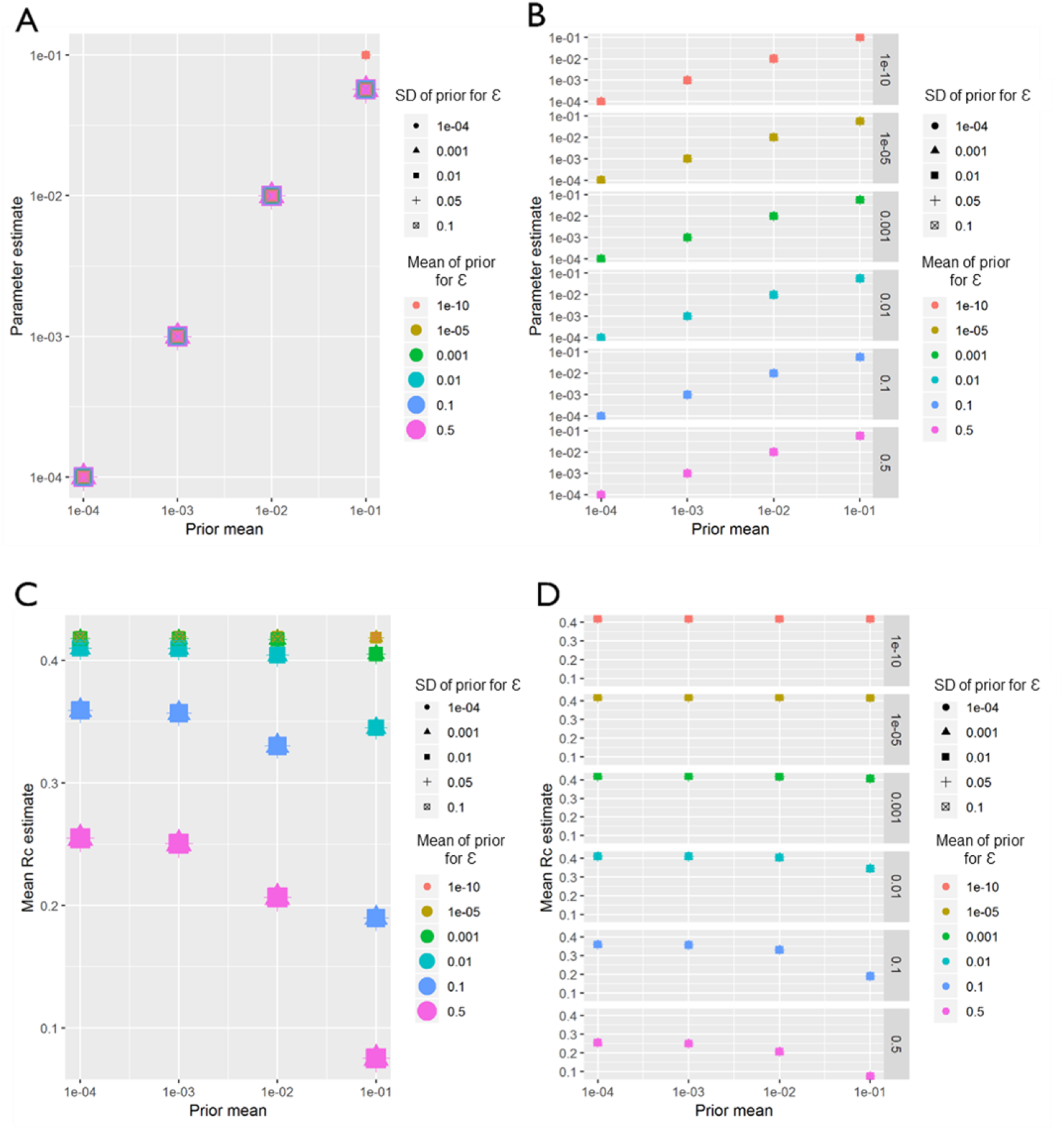
Eswatini sensitivity analysis. Sensitivity analysis showing the impact of varying the prior means for Eswatini. Sensitivity analysis showing the impact of varying the prior mean for the distance kernel shaping parameter, β. The different colours and shapes represent different means and standard deviations respectively of the normally-distributed prior of epsilon, ε, which represents shapes represent different hazards of infection by an external, unobserved source. For A-D, the x-axis represents the prior mean used for β. A) the y-axis shows the maximum a posteriori parameter estimate for the parameter β. B) shows the same results, stratified by the prior mean of ε for clarity. C) Shows the impact of priors for β and ε on the mean Rc estimate, and again D) shows the same result, stratified by the prior mean of ε.

For both *P. vivax* and *P. falciparum* datasets from China, within the parameter range explored in the sensitivity analysis, regardless of how informative the prior was for either β, the distance shaping function, or ε, the epsilon edge, β was always estimated as whatever the mean of the prior was set as (**Figures 11 and 12**), suggesting a lack of identifiability or information within the data. When estimating *R*_*c*_, and interesting interacting effect of ε (missing or unobserved infections) and β (distance) was seen. When β is low, although lower values of ε produce slightly higher mean *R*_*c*_ values, the difference in *R*_*c*_ estimates with varying prior values for ε is much smaller than when β is a higher value. In other words, when the prior for ε is low, 1e-10, *R*_*c*_ estimates do not vary as β changes, however when the prior for ε is much higher, then increasing β from 1e-4 to 0.1 reduces *R*_*c*_ estimates (from 0.21 to 0.01 for *P. vivax)*.

**Figure 11:**
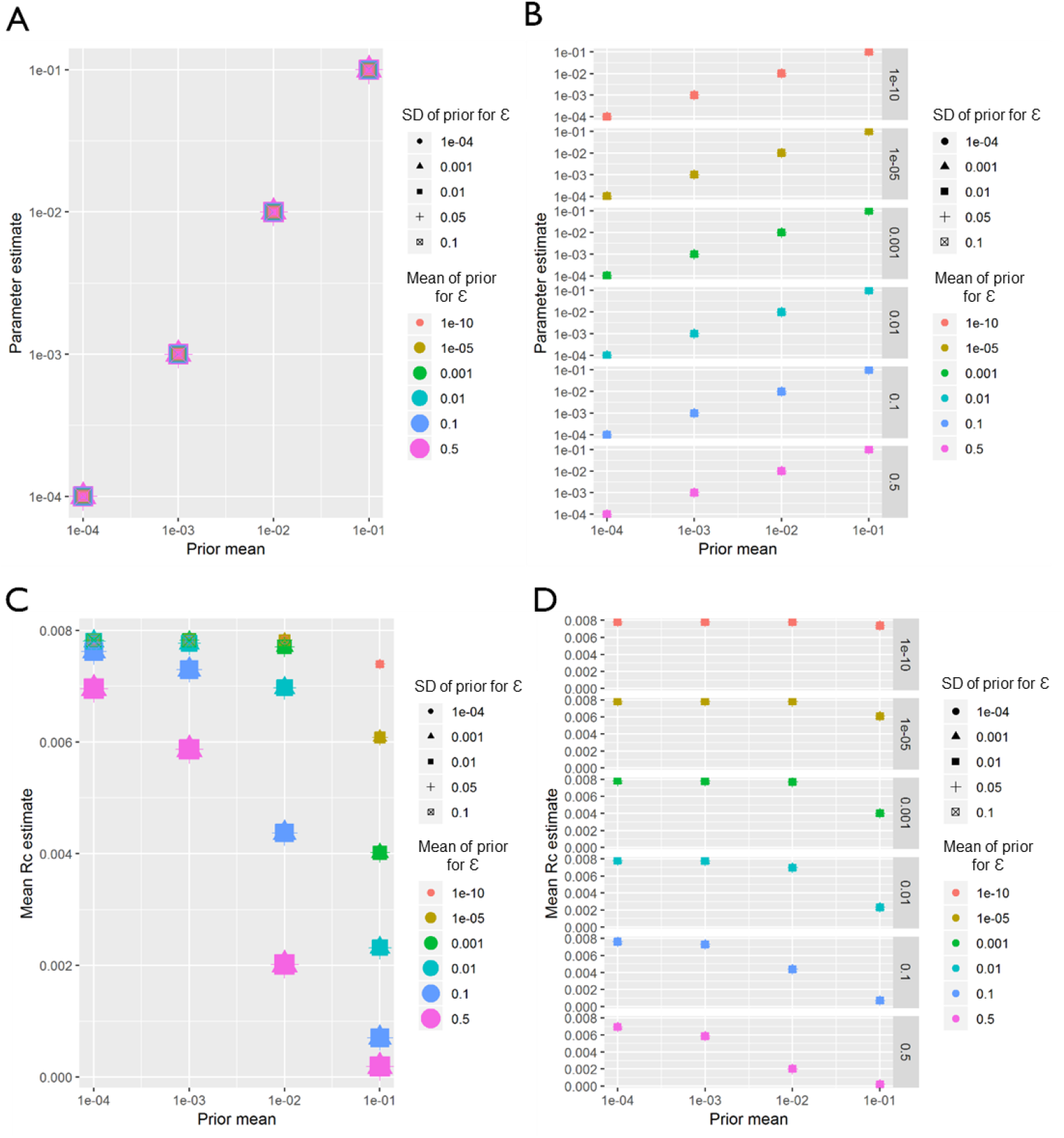
China (P. falciparum) Sensitivity Analysis. Sensitivity analysis showing the impact of varying the prior means for P. falciparum in China. Sensitivity analysis showing the impact of varying the prior means for Eswatini. Sensitivity analysis showing the impact of varying the prior mean for the distance kernel shaping parameter, β. The different colours and shapes represent different means and standard deviations respectively of the normally-distributed prior of epsilon, ε, which represents shapes represent different hazards of infection by an external, unobserved source. For A-D, the x-axis represents the prior mean used for β. A) the y-axis shows the maximum a posteriori parameter estimate for the parameter β. B) shows the same results, stratified by the prior mean of ε for clarity. C) Shows the impact of priors for β and ε on the mean Rc estimate, and again D) shows the same result, stratified by the prior mean of ε.

**Figure 12:**
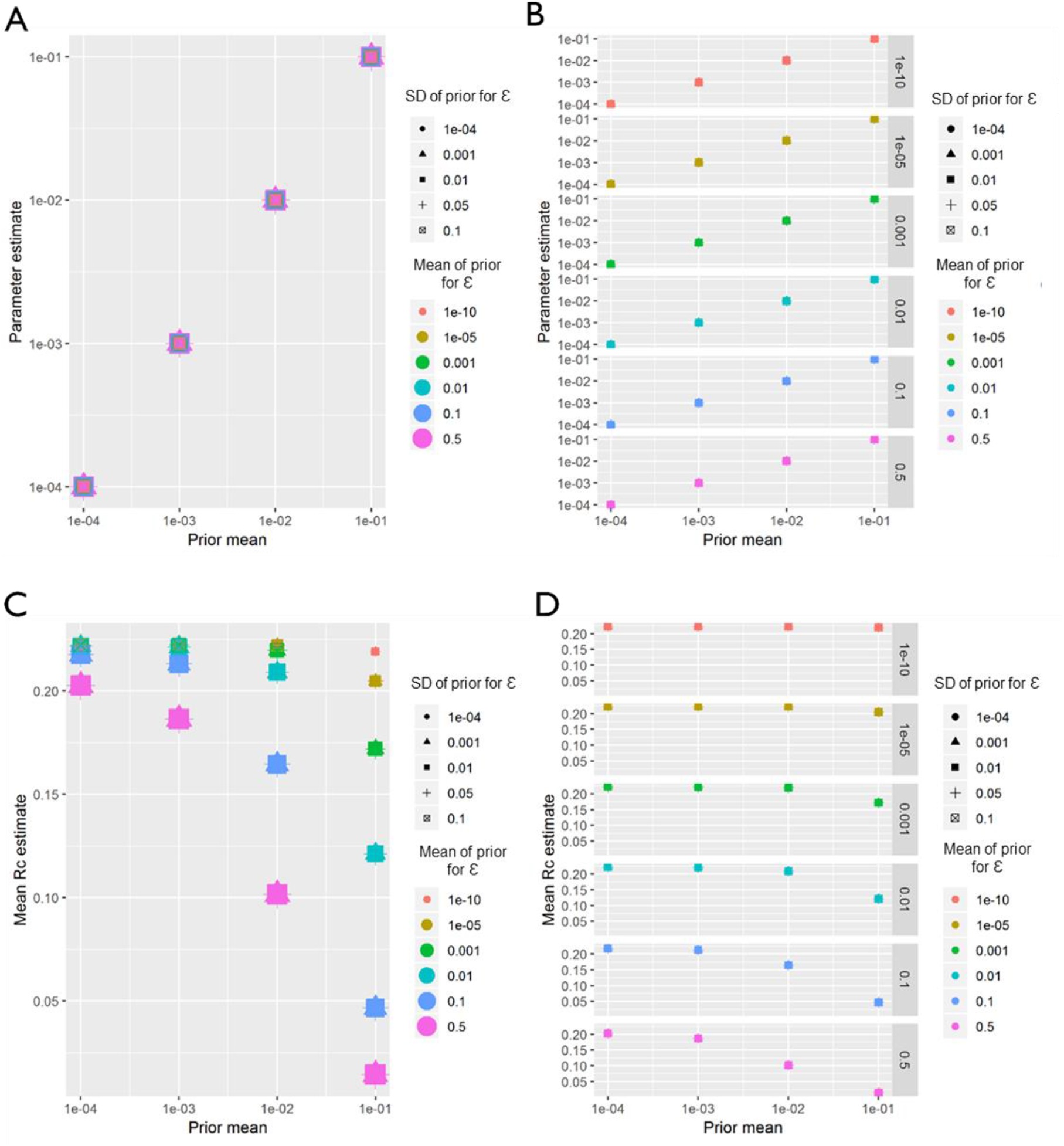
China (P. vivax) Sensitivity Analysis. Sensitivity analysis showing the impact of varying the prior means for P. vivax in China. Sensitivity analysis showing the impact of varying the prior means for Eswatini. Sensitivity analysis showing the impact of varying the prior mean for the distance kernel shaping parameter, β. The different colours and shapes represent different means and standard deviations respectively of the normally-distributed prior of epsilon, ε, which represents shapes represent different hazards of infection by an external, unobserved source. For A-D, the x-axis represents the prior mean used for β. A) the y-axis shows the maximum a posteriori parameter estimate for the parameter β. B) shows the same results, stratified by the prior mean of ε for clarity. C) Shows the impact of priors for β and ε on the mean Rc estimate, and again D) shows the same result, stratified by the prior mean of ε.

## Discussion

We developed an approach to estimate malaria transmission network properties, which allows the flexible integration of distance metrics, such as Euclidian distances or travel times, with temporal information within a single inference framework. Twelve scenarios and corresponding parameter values were defined which represented a) varying likelihoods of transmission over different distances and b) varying likelihoods of missing infections (as well as high and low confidence in this estimate). These scenarios were applied to four individual level datasets from malaria eliminating contexts and using two different spatial kernels. The estimated *R*_*c*_ values, their spatial and temporal distribution and the ΔAICc/Akaike weights for each model were compared alongside a time only model. These results suggest that including spatial information improved models as measured by AIC, compared to time only results. The prior values for both the distance function and epsilon value have very strong impacts on the estimated *R*_*c*_, although relative temporal trends tend to stay consistent.

For all datasets considered, all model versions which used geographic information had lower ΔAICc values than the time only model. Based on the Akaike Weights and ΔAICc values for each model, large differences in ΔAICc were seen between different scenarios. Scenarios 9 and 11 produced the lowest ΔAICc values. These were parameterisations which penalised long range transmission the least where and the prior on epsilon edges was only weakly informative. These parameterisations also return much lower reproduction numbers than using time alone.

Exponential Kernels consistently outperformed Gaussian kernels as measured by ΔAICc. Although classic models of dispersion are as a diffusion process with Gaussian displacement, more leptokurtic or “fatter-tailed” probability distributions, where more of the probability density is concentrated in the tails of the function, are often found to better represent empirical dispersal patterns than traditional Gaussian kernels ^12^. This “fatter-tail” in the exponential can be seen in Figures 13 - 15.

**Figure 13:**
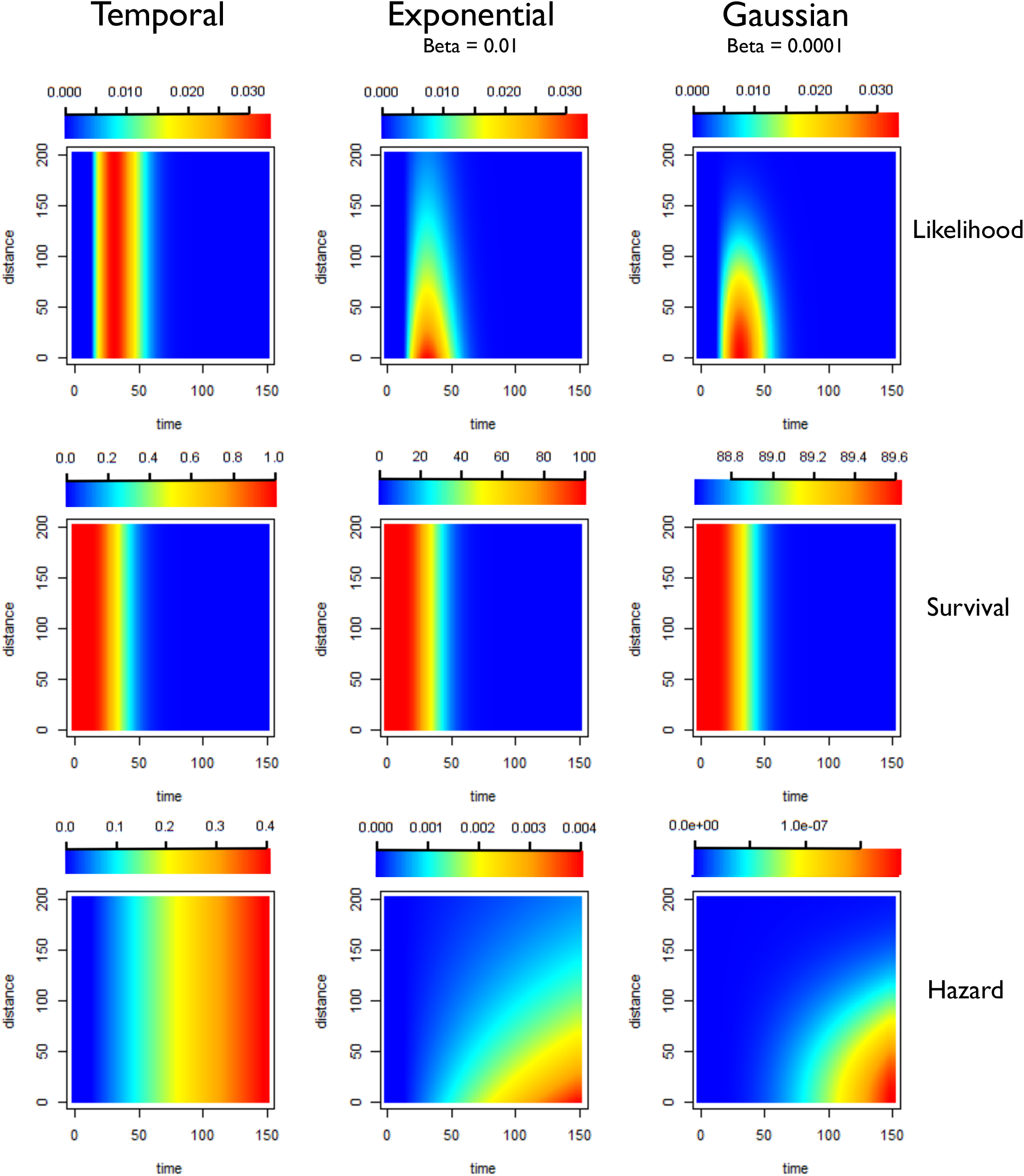
Illustration of likelihoods, hazards and survivals for less restrictive kernels (longer range human movement likely). Plots showing how the pairwise likelihoods, survivals and hazards vary with time and distance under different model structures. The first row of plots shows the pairwise likelihoods, the second row shows the pairwise survival and the third row shows the pairwise hazard values for different combinations of distance (in kilometres) and time between symptom onset (days). The first column shows the results for a time-only version of the algorithm. The second column shows results for an exponential kernel and the third column shows results for a Gaussian kernel. In this example less restrictive values for beta, the shaping parameter for the distance kernels have been chosen, representing a context where there is more long-range movement of parasites.

**Figure 14:**
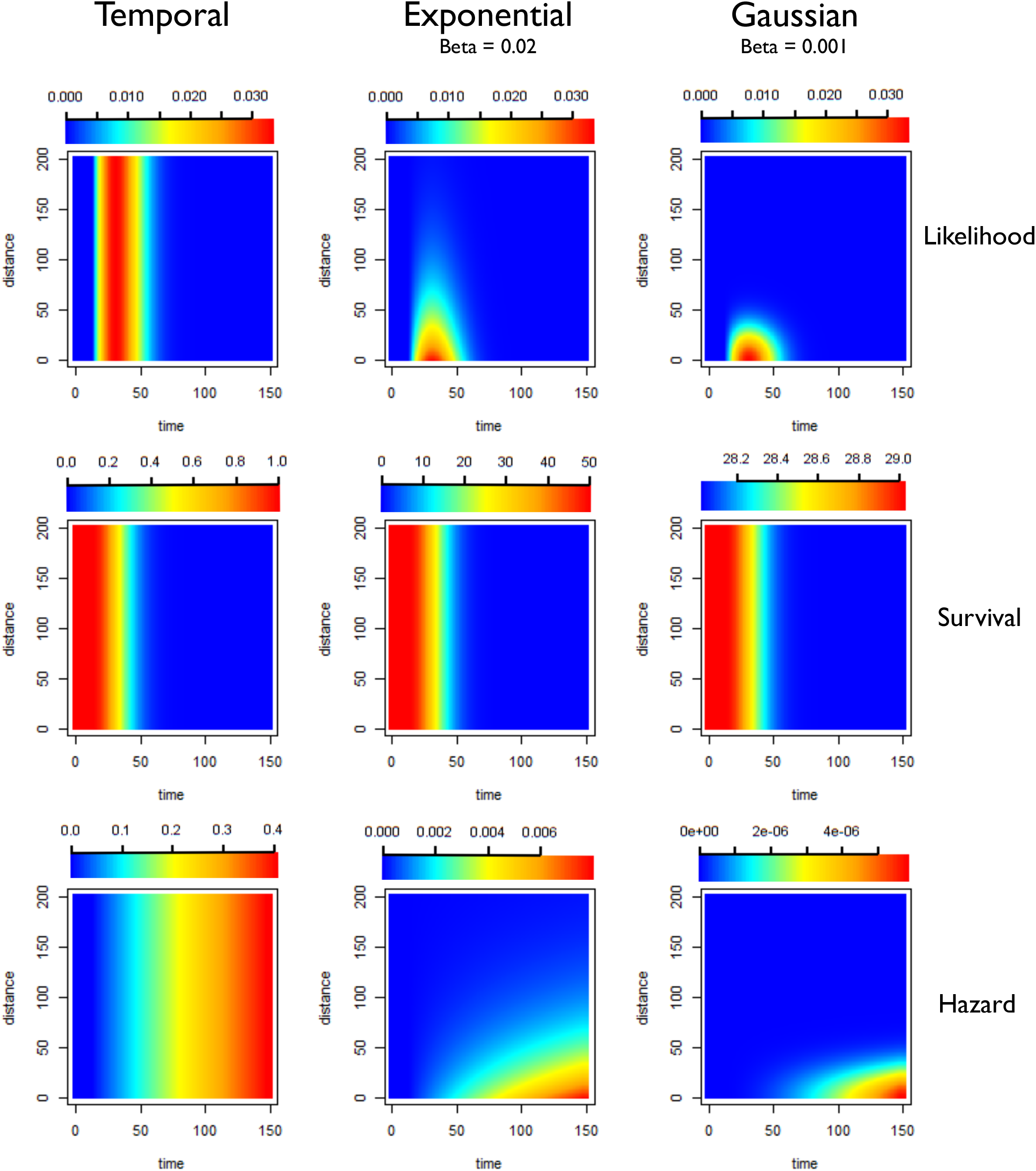
Illustration of likelihoods, hazards and survivals for moderately restrictive kernels (moderate human movement, most movement under 50km). Plots showing how the pairwise likelihoods, survivals and hazards vary with time and distance under different model structures. The first row of plots shows the pairwise likelihoods, the second row shows the pairwise survival and the third row shows the pairwise hazard values for different combinations of distance (in kilometres) and time between symptom onset (days). The first column shows the results for a time-only version of the algorithm. The second column shows results for an exponential kernel and the third column shows results for a Gaussian kernel. In this example values for beta, the shaping parameter for the distance kernels have been chosen to represent a context where there is more some movement of parasites, but where little movement is expected beyond 50-75km. The likelihood for the Gaussian Kernel is more concentrated, which could represent shorter range movement e.g. commutes, whereas the Exponential has a longer tail so could represent a mixture of short and longer range parasite movement.

**Figure 15:**
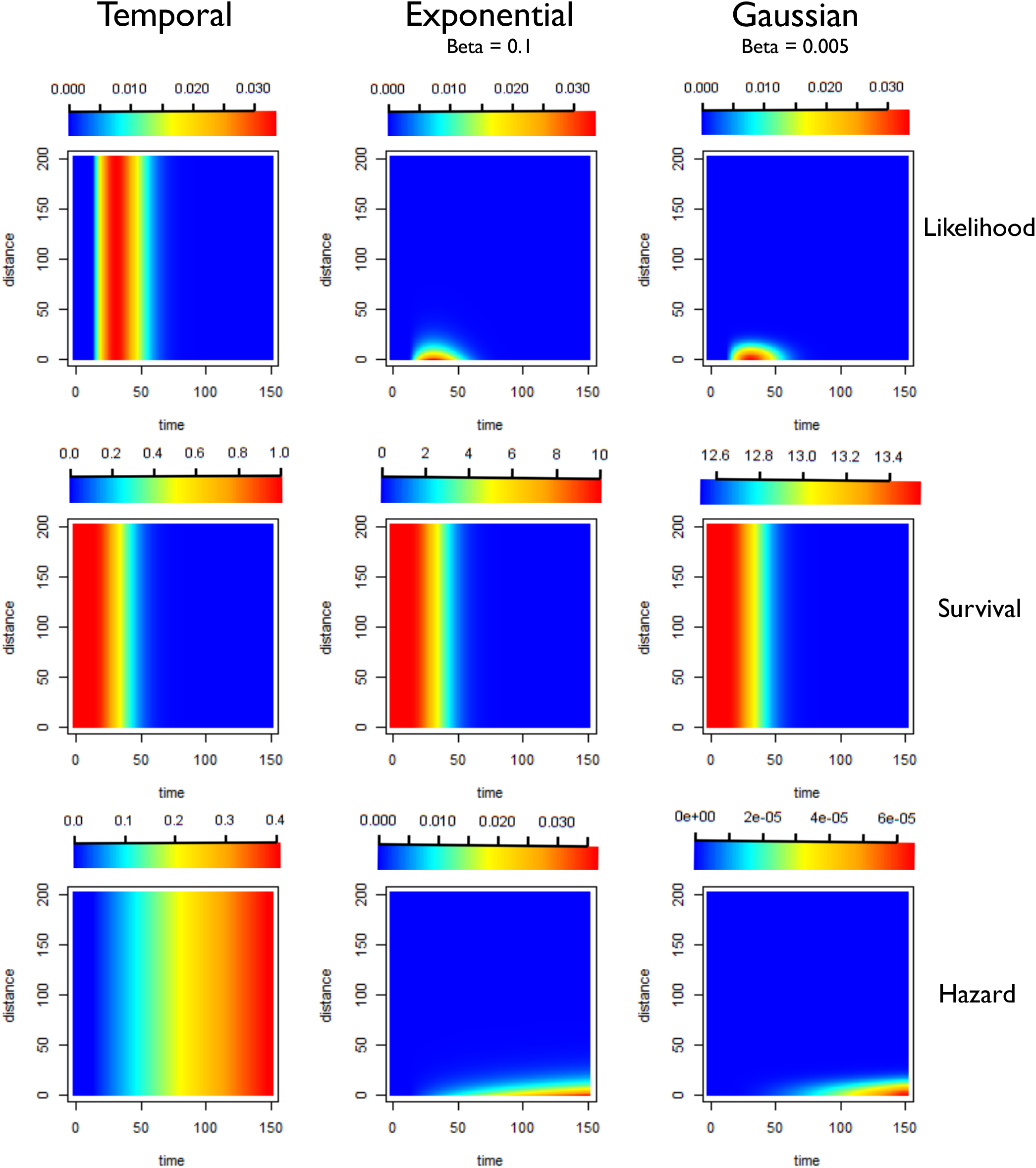
Illustration of likelihoods, hazards and survivals for highly restrictive kernels (Human movement unlikely, most movement under 10km). Plots showing how the pairwise likelihoods, survivals and hazards vary with time and distance under different model structures. The first row of plots shows the pairwise likelihoods, the second row shows the pairwise survival and the third row shows the pairwise hazard values for different combinations of distance (in kilometres) and time between symptom onset (days). The first column shows the results for a time-only version of the algorithm. The second column shows results for an exponential kernel and the third column shows results for a Gaussian kernel. In this example more restrictive values for beta, the shaping parameter for the distance kernels have been chosen, representing a context where there is very little movement of parasites, with very little movement beyond 10-20km expected.

However, there are many limitations to using ΔAICc in model comparison, particularly when estimation of some of the parameters are being carried out within a Bayesian context. We do not fix *α_ij_* nor do we fix epsilon, but we do define priors and maximise the posterior rather than the log likelihood. Therefore, we are comparing negative log likelihoods from a maximised posterior, meaning we are not considering the information included in the prior. In addition, many *α_ij_* values shrink to zero, however are still counted as parameters in the AIC estimation. Therefore, there is no recognition of which versions of the model produce fewer non-zero parameters. Whilst this difference in AIC is interesting to note, I would argue the broader trends in how *R*_*c*_ varies over time and space with different assumptions about both the spatial kernel and the number of unobserved sources of infection are more important to consider.

An interesting pattern which was noted across scenarios and across datasets was how including spatial information in the likelihood tended to increase the seasonality of temporal patterns in reproduction numbers and reduced noise in the temporal distribution of reproduction numbers. This could be suggestive of importation events leading to localised infections. Scenario 4 is also an interesting set of assumptions to consider as it assumes cases generally only infect cases near them and that unobserved cases of infection are unlikely. Under this assumption foci of infection are very clear and clear “sources” of infection.

The results of the sensitivity analysis reveal interesting differences between the different datasets and contexts contained in this dataset. For both El Salvador and Eswatini, which are both small countries (El Salvador has an area of 21,041 km^2^ and Eswatini 17,364 km^2^), at higher mean priors for β, the model converged on an estimate for β which was informed by the data. This was not the case for the dataset from China, which represents a much larger area geographically and where dynamics are likely to be strongly driven by importation. Given that for the kernels we used in this analysis, increasing values of β lead to more restrictive assumptions about the scale of transmission, perhaps this difference is due to the different spatial scales at which the analysis was being carried out.

There are several limitations to this approach and analysis. Firstly, there is a potential lack of identifiability between ε, the epsilon edge, and β, the shaping parameter of the spatial kernel. To give an intuitive example, say two cases occurred 50km from each other in space within a reasonable timeframe of symptom onset times for transmission to have occurred. Without strong prior information about what the spatial kernel may be, and/or how likely cases are to have an external source of infection, it is not clear whether these cases are linked by transmission (and there is some human travel/parasite movement, modelled by a less restrictive spatial kernel) or whether there are unobserved source(s) of infection leading to both cases. This is also exemplified in the results of the sensitivity analysis, where the mean of the prior for beta strongly shapes the final estimate of beta, and the epsilon value also shapes beta.

In the absence of reliable information about either of these values, strong assumptions must be made about either/both the likelihood of cases being infected by unobserved sources of infection and the relationship between distance and. Similar approaches ^13^ recommend fixing the kernel shaping parameter, and indeed approaches from others have also noted problems with unconstrained distance kernels in space-time diffusion modelling (Swapnil Mishra, personal correspondence). One potential way to address this is divide epsilon edge by the distance parameter 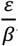, thereby linking the β two parameters and thereby penalising increases in *β*.

Indeed, for similar approaches analysing the diffusion of twitter hashtags, it was recommended to fix the parameter beta, and the authors acknowledged potential challenges in estimating this parameter. Whilst the temporal aspect is not fixed, I view the utility in this method in excluding or penalising improbable transmission links between far away cases, rather than as a way of trying to determine what the spatial relationship between cases is for malaria transmission, or determining the relative contribution of space to malaria transmission.

An additional approach which could alleviate this problem is to collect internal travel history as part of surveillance in future data collection efforts. This may help tease apart the relationship between space and transmission. There also may be regions where there is more information to parameterise both the spatial scales of transmission and the likelihood of cases being unobserved (for example through looking at reporting rates, rates of relapse in the case of *P. vivax*, and prevalence of asymptomatic infection).

Secondly, our approach was designed for application to near elimination and elimination settings, where surveillance and case management is very strong, numbers of cases are small, and therefore there is less overlap in potential infector/infectees, and changes in transmission are more apparent. If applying these approaches to contexts which are less far along the journey to elimination, the issue of identifiability may be even more of an issue as one cannot reasonably assume/fix epsilon edges to be a very small number. Asymptomatic infection will likely be more important to consider, more sophisticated methods to deal with missing cases will be required. There also will likely be a weaker signal in space and time, which may require the integration of additional information such as genetic distance. There also will be a transmission level above which these methods will no longer be useful, although we do not know what this exact level is.

Finally, due to there being no “ground truth” it is hard to rigorously compare model performance. ΔAICc and Akaike Weights are standard, however as mentioned previously, there are important limitations in using these metrics for model comparison. A useful future step would be to analyse simulated line-lists which are spatially explicit to investigate the impact of varying parameter values and the interaction between the shaping parameter of the spatial kernel and epsilon. Spatially explicit simulations may also reveal how tolerant the method is to missingness.

Currently, missing cases are dealt with in a relatively simple way, under the assumption that in the elimination settings used here, surveillance and control have been strong for an extended period of time as to ensure small case numbers and low prevalence of asymptomatic parasitaemia, and that the contribution of missing cases is small enough to be represented as a competing hazard. However, if missingness was biased, it is not clear how strongly this would affect results. Further simulations which model different forms of missing data/sampling schemes would be useful to reveal the potential impact of non-random missing data. These simulations could also model different sources of unobserved infection – for example missing cases caused by relapse of dormant *P. vivax*, unreported cases or asymptomatic infection.

Many methods used to model and represent space and mobility have not been tested here due to the issues of identifiability seen even in simple models of space. Gravity, radiation^14^, and friction surfaces^15^ are all potentially useful models of how space may affect the likelihood of transmission. As mosquitoes have a limited range and lifespan, developing better data and models of human movement, and how it varies in different cultural contexts and between different demographic groups, will provide useful information to appropriately parameterise and design the spatial component of the model.

Although the prior for the shaping parameter of the serial interval was selected under the assumption that the majority of cases are treated in a timely manner, In this analysis we have not explicitly utilised information about the time and location of treatment, although this is available in some contexts. This may be useful information to constrain the potential time window of infection occurring, as detailed information about infectivity and gametocyte carriage following treatment with anti-malarials is available^14^, although sub-optimal dosage, compliance and resistance have been associated with differing outcomes and therefore having additional information about treatment and prevalence of resistance would also be useful. Another avenue for future work would be to adapt the approach to incorporate further sources of information, such as genetic markers of similarity between parasites. For our approach to be useful in contexts which are not at or within a few years of elimination, incorporation of additional information into the inference framework will be required. This could be carried out either directly by incorporating an additional term or function in the likelihood or indirectly through informing the value of parameters and allowing them to vary between individuals. Previous work within the machine learning and network analysis community has successfully integrated diverse sources of information about texts such as language and similarity of context into very similar algorithms to the one presented here ^13^.

## Conclusion

Increasingly, line-list data contain spatial and other forms of information. Developing rigorous approaches to leverage the information contained within these diverse datasets will increasingly be useful in malaria surveillance and epidemiology ^3,5,17^ and developing a framework which flexibly takes on different forms of data within an integrated inference framework is a key aspect of this. There may be more useful information contained in genetic, and or travel, mobility data. However, as we have seen there can be issues of identifiability, which becomes increasingly relevant when there is not enough data available about key parameters in the model. Finding ways for leveraging multiple datasets, understanding their relationships, how they can enhance info contained in others, or used to build consensus is important.

We developed and tested an algorithm which flexibly allows the incorporation of distance or adjacency matrices describing the distance or connectivity between cases. This was applied to individual malaria case data from four eliminating and very low transmission contexts and a detailed sensitivity analysis was carried out. The results of these analyses suggest that including space improves model performance as measured by ΔAICc, and that, for the contexts considered here, the best performing models produce lower reproduction estimates than using temporal information only, likely in part due to estimating more unobserved sources of infection. However, this conclusion would be strengthened by more in-depth simulation studies. The approach presented here could be adapted to many different datasets and contexts, however issues of identifiability must be considered. The utility of this approach would be strengthened with further development of the methods of modelling unobserved sources of infection. Our results also make it clear that in many contexts that additional information sources may be required such as genetic or serological data.

## Methods

### Data

#### The Kingdom of Eswatini

This dataset, previously analysed by Reiner and colleagues^18^ captures malaria cases recorded by the National Malaria Elimination Programme in the Kingdom of Eswatini (formally known as Swaziland) between January 2010 and June 2014. For each case detected during this time (N= 1373), case investigation was carried out. For each case the following were collected: GPS coordinates of household location, demographic information (age, occupation and sex), use of malaria prevention interventions such as long-lasting insecticide treated bednets (LLINs), and date of symptom onset, diagnosis and treatment, as well as travel history. Based on travel history cases were defined as locally acquired, imported. For a small number of cases (N=58) the local/imported status was determined “unknown”. For the purposes of this analysis, these cases were treated the same as local cases, i.e. they were assumed to have potentially been infected by other cases in the dataset and/or been infectors themselves.

#### China

This dataset consists of individual-level case data for all confirmed and probable cases reported in China between 2011 and 2016 ^9,19^ (Table 4 and Table 5). The data consist of an individual identifier, date of symptom onset, date of diagnosis and date of treatment, as well as the geolocated address of residence and health facility. If the suspected location of infection was in China and not in the same district, then the presumed location of infection was also included in the dataset. Demographic information such as age and sex were also collected. For the analysis, data were separated into *P. falciparum* and *P. vivax* cases. *P. malariae* (N=252) and *P. ovale* (N=822) were reported but excluded from the analysis due to the lower public health concern of these species.

**Table 4:** Cases by diagnosis type (probable and confirmed) and species across China.

|  | Mixed infection | *P. falciparum* | *P. malariae* | *P. ovale* | *P. vivax* | Untyped |
| --- | --- | --- | --- | --- | --- | --- |
| **Confirmed** | 260 | 11830 | 252 | 822 | 6631 | 87 |
| **Probable** | 0 | 176 | 0 | 0 | 693 | 311 |

**Table 5:** Cases by imported/local status and species across China.

|  | Mixed infection | *P. falciparum* | *P. malariae* | *P. ovale* | *P. vivax* | Untyped |
| --- | --- | --- | --- | --- | --- | --- |
| **Local** | 5 | 92 | 4 | 1 | 1711 | 95 |
| **Imported** | 255 | 11914 | 248 | 821 | 5613 | 303 |

#### El Salvador

This dataset consists of all confirmed cases of malaria recorded by the Ministry of Health in El Salvador between 2010 and the first two months of 2016^11^ (N= 91 cases, of which 30 imported, 6 *P. falciparum*, 85 *P. vivax*). For each case, the date of symptom onset was recorded. Residential address was available for all but two cases. For these cases, the location was available at the *municipio*, or municipality level, and the coordinates of the centroid of the municipality (which for both were cities) were used as the geo-location. Two cases had addresses listed outside of El Salvador, both of which were located in Guatemala. All cases within El Salvador with full addresses (N=85) were georeferenced by latitude and longitude to *caserío/lotificación* level, which is approximately neighbourhood or hamlet level.

### Transmission model specifics

In order to incorporate pairwise distance metrics, we extended our previously published algorithm applied to Yunnan Province, China^9^ by introducing a second function, *f*_*2*_, which describes the relationship between space (or distance of any kind) and likelihood of transmission. An appropriate function such as a Gaussian kernel is defined and the parameter(s) shaping that distribution, β, are either fixed, or given a prior distribution and estimated from the data. Multiplied, together, this returns a single function:

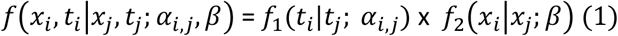

Determined by times *t*, spatial locations *x*, transmission rates α, spatial parameter(s) β.

As before, the hazard is defined as the pairwise likelihood divided by the survival term:

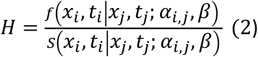

To derive the survival function, one integrates across all distances and times as follows:

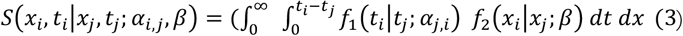

The specific functions used in *f*_1_(*t*_*i*_|*t*_*j*_; *α*_*i,j*_) and *f*_*2*_(*x*_*i*_|*x*_*j*_; *β*) will have large impacts on the outcomes of results and therefore the assumptions inherent in these choices must be made explicit and linked to the mechanisms of transmission.

To illustrate this approach by applying to several malaria line-lists, we used a shifted Rayleigh distribution to model serial interval distributions, *f*_1_(*t*_*i*_|*t*_*j*_; *α*_*i,j*_). For the second part of the likelihood which model the relationship between space and the likelihood of transmission *f*_*2*_(*x*_*i*_|*x*_*j*_; *β*), Gaussian and Exponential diffusion kernels were used (Table 6).

**Table 6:**
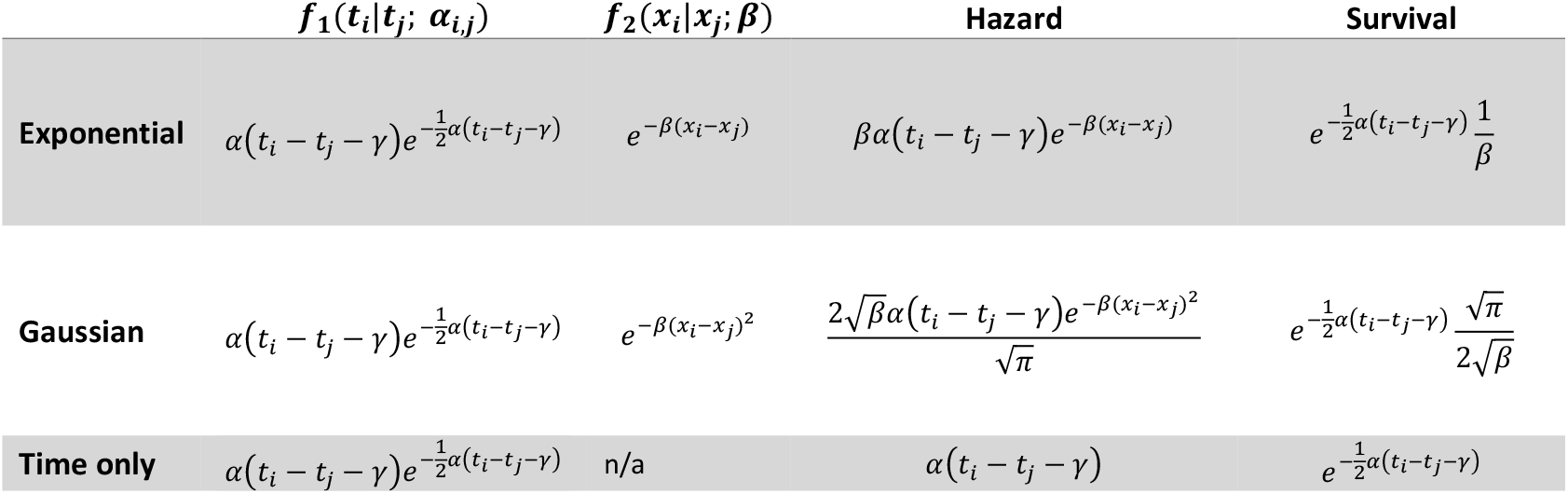
Equations for f1, f2, hazard and survival for time -only, Gaussian and Exponential spatial kernels.

Using a shifted Rayleigh distribution and an exponential kernel the pairwise likelihood of a case showing symptoms at *t*_*i*_ and at residence location *x*_*i*_ being infected by a case showing symptoms at time *t*_*j*_ and at residence location *x*_*j*_, becomes

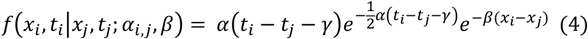

The survival term simplifies to:

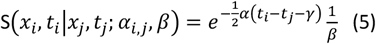

And the hazard simplifies to:

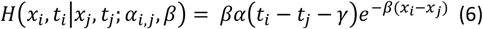

For the Gaussian function, the pairwise likelihood of a case showing symptoms at *t*_*i*_ and at residence location *x*_*i*_ being infected by a case showing symptoms at time *t*_*j*_ and at residence location *x*_*j*_ is

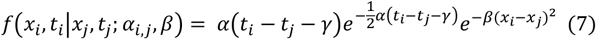

The survival term is again determined by integrating the likelihood over all potential infection times and all distances

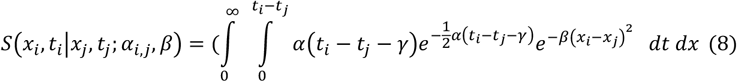

Integrating over time returns

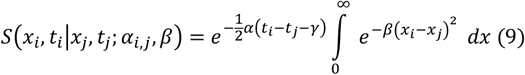

Integrating over all distances gives

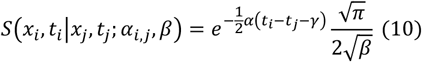

Following equation 10, the hazard is equivalent to

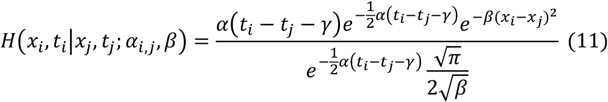

Which simplifies to

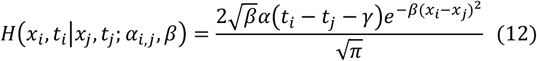

### Modelling missing cases using *ε* edges

The vast majority of disease surveillance and outbreak response datasets will not be able to capture all cases due to asymptomatic infection, underreporting and movement of people in/out of the surveillance area. Therefore, it is important to consider the impact of missing information on results and identify potential missing sources of infection. We use Epsilon edges, *ϵ*_*i*_, to identify potential sources of infection. Here, each hazard is estimated as a further competing edge of transmission from an unobserved source, *H*_*0*_(*ϵ*_*i*_). Depending on assumptions for the likelihood and extent of unobserved infection sources, the epsilon edge value can be set to a high or low value. When high, we assume high amounts of unobserved infection and unless two cases have a very high likelihood of being linked, we assume the case was from an unobserved source. When low, we assume little missing data and so cases are only linked to an outside source if they are very unlikely to be linked to an observed candidate infector.

Adding epsilon, ε, as a competing hazard and survival returns:

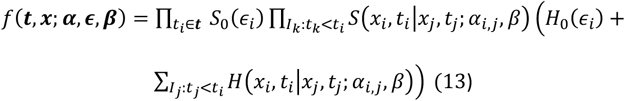

The objective function is then:

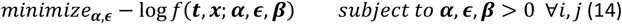

Because this was carried out within a Bayesian framework the log posterior was maximised to obtain the maximum-a-posteriori estimates.

The algorithm was written in TensorFlow, implemented in R via the *rTensorflow* package. A prior probability was defined for the parameter shaping the serial interval of malaria, informed by previous characterisations of the serial interval of malaria ^20^. Because data about how likely the cases were to have moved long distances or the likelihood of a case has been infected by an unobserved source of infection were not available for the contexts explored here, several different parameterisations of the model were used to represent different scenarios (Table 1) and a detailed sensitivity analysis was carried out (Table 3). The versions of the model which are described in **Table 1** and **Figures 13-15** represent different patterns of human/parasite movement, ranging from a context where there may be small amounts of movement (almost all under 10km) to moderate amounts of movement/travel(almost all under 50km) to a less restrictive parameterisation, where near cases were more likely but far away cases were not completely excluded. We applied different versions of the algorithm, as well as temporal-only algorithm to these datasets to explore the impact of different assumptions about the impact of space on estimated *R*_*c*_ values and their variation over time and space. We also evaluated the performance of each approach by comparing differences in the second order AIC (ΔAICc), and the corresponding Akaike Weights.

Twelve scenarios (**Table 1**) were considered when defining parameters for each dataset. These scenarios consider three different levels of likelihood of transmission in relationship to Euclidian distance (due to the limited range of mosquito travel, this is considered in the context of human mobility), which was defined for both exponential and Gaussian kernels. These are illustrated in **Figures 13-15**. Then the values for epsilon were set at 0.001 and 0.1, representing different levels of missing cases likely. This can be interpreted as the chance of a case having an unobserved source of infection. For example, 0.1 would represent P(unobserved source of infection) = 0.1.

The timeseries of *R*_*c*_ and its spatial patterns were illustrated for each dataset and parameter combination and compared to the results of the time-only version of the algorithm. The results were also mapped to compare how spatial patterns in *R*_*c*_ were affected by assumptions about space and unobserved infections.

In order to compare models quantifiably, the second order Akaike Information Criterion (AICc) was calculated using the equation 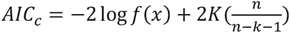,where *f(x)* is the model likelihood, K is the number of parameters estimated and n is the sample size of the data used to fit the parameters. The AIC^21^ is used in model comparison, by creating a comparison of negative log likelihood that penalises increases in model parameters, to prevent overfitting. AICc is recommended for use with smaller datasets with larger numbers of parameters, and as the sample size *n* increases AICc converges to AIC^10^. The differences in AICc for each model, known as ΔAICc, were calculated to compare models. Typically, a ΔAICc of greater than 10 is considered strong evidence that that model performs worse than the model it is being compared to.

In addition, Akaike Weights were calculated, which are a measure of the relative likelihood of a model compared to the others considered. Akaike weights are determined by taking the normalised relative likelihood of a model which is *exp*(−*0*.5 * Δ*AICc score*), and then dividing by the sum of these values across all models to obtain a normalised result.

### Sensitivity analysis and comparison of prior choice on estimated results

In the scenario analysis above the distance shaping parameter is fixed. However due to the uncertainties in the relationship between distance and likelihood of transmission, in many contexts it may be useful to estimate *β*. To explore the relationship between the estimated epsilon edges, *ϵ*, and estimated shaping parameter, *β*, for the distance function. a detailed sensitivity analysis was carried out to explore the impact of a) prior choice for *ϵ* d) prior choice for *β* on both the maxmum-a-posteriori estimates for *β* and the estimated mean *R*_*c*_.

To consider the effect of varying parameter values and explore their interactions, a range of distance and epsilon edge priors were considered. A truncated normal prior was used for both parameters, and the mean and standard deviation were varied. For *ϵ* the mean was varied between 1e-10 and 0.5, and the standard deviation was varied between 0.0001 and 0.1. For *β*, the mean for a Gaussian Kernel was varied between 0.00001 and 0.01 and for an exponential kernel the means considered ranged between 0.0001 and 0.1. For both the standard deviations varied between 0.0001 and 0.1 (Table). Every possible combination of the parameters were run for each dataset and both Gaussian and exponential spatial kernels, giving a total of 2400 parameter combinations tested per kernel, per dataset.

## Data Availability

The underlying patient line-list data used in the study from China are available from The Chinese Center For Disease Control and Prevention, 155 Changbai Road Changping District, Beijing 102206,China. Tel: +86-10-58900240, 58900216 or from Dr Junling Sun. She is the Head of the Branch of Parasitic Disease, Division of Infectious Disease, China CDC.
The line list data from El Salvador are not publicly available because they are nationally-owned data therefore the authors do not have the permission to host them but are available from authors upon reasonable request and with permission of the Ministry of Health, El Salvador (MINSAL). 
Rc estimates for time-only model and code to generate them are available from GitHub.

https://journals.plos.org/ploscompbiol/article?id=10.1371/journal.pcbi.1007707

https://github.com/IzzyRou/estimates-Rc-China

## Supplementary Information and Figure Legends

### Supplementary Information

**Supplementary Table 1:** Full results of ΔAICc and Akaike Weights for each scenario, dataset and spatial kernel considered

